# Genomic rare variant mechanisms for congenital cardiac laterality defect: A digenic model approach

**DOI:** 10.1101/2024.11.19.24317385

**Authors:** Archana Rai, Jonathan Klonowski, Bo Yuan, Karen J. Coveler, Zain Dardas, Iman Egab, Jiaoyang Xu, Philip J. Lupo, A. J. Agopian, Dennis Kostka, Cecilia W. Lo, Stephen (Song) Yi, Bruce D. Gelb, Christine E. Seidman, Eric Boerwinkle, Jennifer E. Posey, Richard A. Gibbs, James R. Lupski, Shaine A. Morris, Zeynep Coban-Akdemir

## Abstract

Laterality defects are defined by perturbations in the usual left-right asymmetry of organs. Due to low known genetic etiology of congenital heart disease (CHD) cases (less than 40%), we used a digenic model approach for the identification of contributing variants in known laterality defect genes (N = 115) in the exome/genome sequencing (ES/GS) data from individuals with clinically diagnosed laterality defects. The unsolved ES/GS data were analyzed from three CHD cohorts: Baylor College of Medicine-Genomics Research to Elucidate the Genetics of Rare Diseases (BCM-GREGoR; N = 247 proband ES), Gabriella Miller Kids First Pediatric Research program (Kids First; N = 158 trio GS), and Pediatric Cardiac Genomics Consortium (PCGC; N = 163 trio ES), and trans-heterozygous digenic variants were identified in 2.8% (inherited digenic variants in 0.4%), 8.2%, and 13.5% cases respectively, which was significantly higher as compared to 602 control trios provided by the 1000 Genomes Project *(p =* 0.001, 1.4e-07, and 8.9e-13, respectively). Trans-heterozygous digenic variants were also identified in 0.4%, and 1.4% cases with non-laterality CHD in Kids First and PCGC datasets, respectively, which was not statistically significant as compared to control trios (*p* = 1, and 0.059, respectively). Altogether, in laterality cohorts, 23% of digenic pairs were in the same structural complex of motile cilia. Out of 39 unique digenic pairs in laterality CHD, 29 are more likely to be potential digenic hits as predicted by DiGePred tool. These findings provide further evidence that digenic epistatic interaction can contribute to the complex genetics of laterality defects.

## Introduction

Laterality developmental abnormalities, including congenital cardiac laterality defects, are rare congenital anomalies of embryonic left-right (LR) axis patterning. In humans, they occur with a frequency of ∼1.1 cases per 10,000 live births^1^ and account for ∼3-7% of all congenital heart disease (CHD).^2,3^ They involve a spectrum of disorders that range from D-transposition of the great arteries affecting only the heart to complex conditions including situs inversus totalis (SIT) and heterotaxy (HTX), which affect the entire thorax and abdomen. SIT is characterized by complete, mirror-image reversal of all asymmetrical structures; whereas HTX is defined as having at least one organ discordant along the left-right axis and is traditionally classified into two groups: left atrial isomerism and right atrial isomerism, although some forms of HTX cannot be easily classified into these 2 groups.^4^ The heart exists as a midline linear tube early in cardiogenesis, and undergoes multiple looping stages before reaching its final four-chambered configuration; it is particularly sensitive to LR signaling cues for normal development. Abnormal LR patterning therefore frequently results in complex CHD. Patients with laterality defects complicated by CHD have higher mortality than patients with CHD contemporaries with no laterality defect. A previous study revealed that the postoperative mortality of CHD patients with HTX after surgical treatments was 4.8%, compared with 2.4% for CHD patients who did not have HTX, even when controlling for severe CHD.^5,6^

Laterality defects show extensive heterogeneity in their genetic inheritance (autosomal dominant, autosomal recessive, X-linked, multigenic, etc.), genetic etiology, and phenotypic presentation.^7,8^ However, current clinical genetic testing identifies an underlying genetic cause in less than 40% of cases,^9^ indicating that additional genetic mechanisms and gene contributors remain to be elucidated. Clinically relevant copy number have been identified in 15% to 26% of patients with heterotaxy syndrome.^10,11,12^ While a monogenic cause is not identified in many individual cases, a more complex genetic model could be considered. For instance, digenic inheritance models may contribute to the complex genetics of these defects and others.^13,14^ In the current study, we performed a comprehensive genetic analysis on unsolved exome/genome sequencing (ES/GS) data of individuals with laterality congenital cardiac defects from three CHD cohorts and investigated a digenic model, focusing our analysis on genes with known variant alleles already associated with laterality defects. A conceptual framework of this study is summarized in **Figure 1**. Our findings illustrate the potential contribution of a digenic model of disease, whereby digenic epistatic interaction can contribute to the complex genetics of laterality defects.

**Figure 1:**
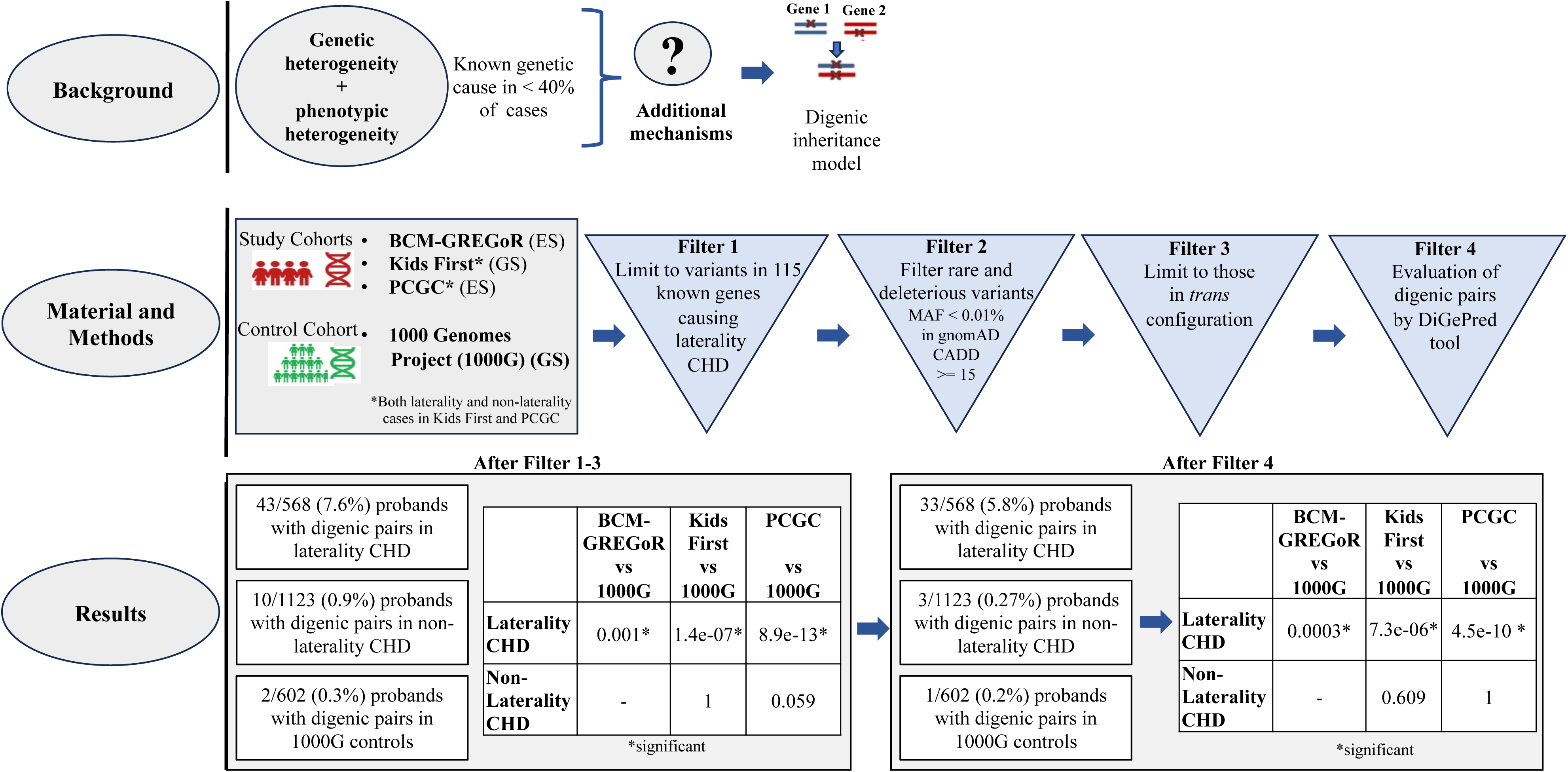
Conceptual framework of the study. Cases with Laterality and non-laterality defects were analyzed for digenic variants in known 115 laterality genes from three cohorts, BCM-GREGoR, Kids First, and PCGC. 602 healthy population from 1000G were used as control cohort. Our analysis shows that the proportion of individuals carrying trans-heterozygous digenic variants differs significantly between control cohorts and laterality defect cohorts (*P* = 0.001, 1.4e-07, and 8.9e-13 in BCM-GREGoR, Kids First, and PCGC vs. 1000G). However, no significant difference was observed between controls and non-laterality cases (*P* = 1, 0.059 in Kids First and PCGC vs. 1000G). 29 out of 39 unique gene pairs identified in three laterality cohorts, were likely to be potential digenic hits based on their pathway similarity, phenotype similarity, co-expression rank, protein-protein interaction distance, and pathway distance (*P* = 2.2e-16). 10 gene pairs are are in same structural complex of motile cilia. ES: exome sequencing; GS: genome sequencing.

### Materials & Methods

#### Study cohorts

The study included laterality defect cases from the three CHD cohorts: Baylor College of Medicine-Genomics Research to Elucidate the Genetics of Rare Diseases (BCM-GREGoR; N = 247 probands; ES), Gabriella Miller Kids First Pediatric Research program (Kids First; N = 158; trio GS), and Pediatric Cardiac Genomics Consortium (PCGC; N = 163; trio ES). BCM-GREGoR cohort included 247 “unsolved” proband-only exomes of individuals with clinically diagnosed laterality defects recruited through Texas Children’s Hospital (TCH) in Houston, TX following the Institutional Review Board of Baylor College of Medicine approved research study protocol (IRB approval number: H-1843) and Baylor Genetics (BG) for clinical exomes (IRB approval number: H-48014). Written informed consent was obtained from all participating individuals including probands and any available family members prior to their enrollment. The Kids First and PCGC programs included unsolved 158 and 163 trios presenting with laterality defects, respectively. In PCGC program, CHD subjects were recruited to the Congenital Heart Disease Genetic Network Study of the Pediatric Cardiac Genomics Consortium (CHD GENES: ClinicalTrials.gov identifier NCT01196182).^15^ The institutional Review Boards of Boston’s Children’s Hospital, Brigham and Women’s Hospital, Great Ormond Street Hospital, Children’s Hospital of Los Angeles, Children’s Hospital of Philadelphia, Columbia University Medical Center, Icahn School of Medicine at Mount Sinai, Rochester School of Medicine and Dentistry, Steven and Alexandra Cohen Children’s Medical Center of New York, and Yale School of Medicine approved the protocols.^15^ All subjects or their parents provided informed consent. This Kids First project represents a subset of the PCGC data, so CHD subjects were recruited as per PCGC study protocol. The PCGC and Kids First datasets may contain overlapping cases, which will be identified and removed from the analysis. We also studied an additional 551 and 572 trios with non-laterality CHD in Kids First and PCGC cohorts, respectively. The distribution of Kids First and PCGC cases in terms of their additional CHD features are given in **Table S1**. All the sample IDs used in this study are research lab identifier. No one outside the research group have access to the sample ID.

### The genes known to be associated with laterality defect

A set of 115 genes with known laterality defects associations was used in this study to identify variants in laterality defect cases in those three cohorts. These 115 genes are known to be associated with LR patterning abnormalities or HTX based on the literature evidence of phenotypic data in humans.^9^ Out of 115 genes, 55 (48%) were ciliary genes, and 60 (52%) were non-ciliary genes **(Table S2**). In 9 genes, variants are known to be associated with autosomal dominant (AD) inheritance, 65 genes with autosomal recessive (AR), three genes (*TTC21B*, *ACOX1,* and *GDF1*) with both AD and AR manner, six genes as X-linked recessive (XLR), one gene (*BCOR*) as X-linked dominant (XLD), one gene (*FLNA*) as both XLD and XLR, and thirty genes are with unknown inheritance pattern.

### Identification of digenic variants among three laterality defect cohorts

Laterality CHD have considerable heterogeneity in their genetic origins and current clinical genetic testing identifies an underlying genetic cause in less than 40% of cases,^9^ indicating that additional mechanisms need to be explored. Therefore, we aimed to examine a digenic model approach by screening each exome/genome of unsolved cases in the three laterality CHD cohorts for variants in 115 known laterality defect genes. To detect potential digenic variants, a stepwise analysis workflow was implemented.

### Sequencing, annotation and variant selection in BCM-GREGoR cohort

For the BCM-GREGoR cohort, ES was performed at the Human Genome Sequencing Center (HGSC) at Baylor College of Medicine (BCM) through the Baylor Hopkins Center for Mendelian Genomics (BHCMG) initiative. The paired-end pre-capture library was constructed using Illumina platform according to the manufacturer’s protocol (Illumina Multiplexing_SamplePrep_Guide_1005361_D) with modifications as described in the BCM-HGSC Illumina Barcoded Paired-End Capture Library Preparation protocol. Paired-end sequencing was performed with the Illumina HiSeq 2000 platform or Illumina NovaSeq 6000 platforms. Reads were aligned to GRCh37/hg19 with BWA-aln (for data generated on HiSeq 2000) or BWA-mem (for data generated on NovaSeq). Sequence analysis was performed with the HGSC Mercury analysis pipeline (https://www.hgsc.bcm.edu/software/mercury),^16,17^ and variant calling was performed using ATLAS2 or xATLAS,^18^ and the Sequence Alignment/Map (SAMtools) suites. Variants were functionally annotated using our in-house developed Cassandra^19^ annotation pipeline which utilizes Annotation of Genetic Variants (ANNOVAR) and additional tools and databases, including gnomAD (https://gnomad.broadinstitute.org), and the Atherosclerosis Risk in Communities database (https://aric.cscc.unc.edu/aric9/).

The selection of variants was based on the identification of missense and protein truncating variants. Rare variants, minor allele frequency (MAF) <0.01%, were prioritized according to frequency in the population databases including the 1000 Genomes Project (1000G), gnomAD v4.0, and our in-house-generated ES database (ES from ∼13,000 individuals) at the BCM-GREGoR. Rare variants with a Combined Annotation Dependent Depletion (CADD)-phred score of >= 15 were included. The analysis was complemented with two gene prioritization strategies to identify a list of rare variants located in known laterality defect genes. In the BCM-GREGoR cohort, we have proband data only. Therefore, further analysis of candidate variants was performed on a family-by-family basis via Sanger sequencing to look for trans-heterozygous pattern. Digenic events were defined as combinations of candidate variants in two known laterality CHD genes co-segregating with disease, i.e., combinations of candidate variants present in affected individuals of each family but not in unaffected individual of the family. Variants should be inherited from the parents — at least one each from the mother and the father. All parents were unaffected, with the exception of one family, in which one of the parents of a female proband was affected.

### Sequencing, annotation and variant selection in Kids First cohort

GS trio data of Kids First CHD cases were obtained from the Database of Genotypes and Phenotypes (dbGaP) study with accession number phs001138.v4.p2. For GS, DNA was fragmented and sequenced to 30x mean coverage on an Illumina HiSeq 10X, generating 150 bp paired-end read and read quality assessment was done by FastQC. Alignment and germline variant calling were performed by CAVATICA workflow (https://cavatica.sbgenomics.com; “Whole Genome Sequencing - BWA + GATK 4.0 (with Metrics),” Revision 41). Reads were aligned to hg38 with BWA-mem, and further, variants were called using HaplotypeCaller after base quality score recalibration (BQSR). Variant quality score recalibration (VQSR) was applied to SNPs and indels using several training files available from the GATK Resource Bundle (hapmap3.3 sites, 1KG_omni2.5, 1KG_snp/indel_high.confidence, and dbsnp_135 files). Variants were functionally annotated using ANNOVAR 2019Oct24 (https://annovar.openbioinformatics.org/)^20^ with the following datasets: refGene, cytoband, exac03, avsnp147, dbnsfp30a, gnomad211_genome, gnomad211_exome, clinvar_20220320, intervar_20180118, esp6500siv2_all, 1000g2015aug_all, 1000g2015aug_afr, 1000g2015aug_eas, and 1000g2015aug_eur. The selection of variants was performed as previously described for the BCM-GREGoR cohort and analyzed for combination of candidate variants in two known laterality defect genes co-segregating with disease. Here, we have the sequencing data from parents as well, so the trans-heterozygous pattern of digenic variants was examined for each family.

### Sequencing, annotation and variant selection in PCGC cohort

ES data of PCGC probands and parents were obtained from dbGaP with accession number phs001735.v2.p1, and phs001194.v2.p2 respectively. Trios were sequenced at the Yale Center for Genome Analysis. DNA was captured with the NimbleGen v2.0 exome capture reagent (Roche) or Nimblegen SeqxCap EZ MedExome Target Enrichment Kit (Roche) and sequenced in paired-end mode (Illumina HiSeq 2000, 75 base paired-end reads). Two independent analysis pipelines were used at Yale University School of Medicine (YSM) and Harvard Medical School (HMS). At each site, reads were independently mapped to theGRCh37/hg19 reference genome using BWA-mem (YSM) and Novoalign (HMS). Further processing was performed by GATK Best Practices workflows and variants were called using HaplotypeCaller. Variants were functionally annotated using ANNOVAR 2019Oct24 using datasets same as in Kids First, and selection of variants was performed as previously described for BCM-GREGoR cohort and looked for combination of candidate variants in two known laterality defect genes co-segregating with disease. Here, we have the sequencing data from parents as well, so each family was analyzed for the presence of trans-heterozygous variants co-segregating with disease.

### Frequency comparison of digenic variants among cohorts

We hypothesized that damaging digenic combinations of alleles in laterality CHD genes should be in *trans* in affected individuals and should not coexist in *trans* in the healthy populations or should show lower frequencies than expected by chance. Additionally, we hypothesized that the prevalence of subjects with trans-heterozygous digenic variants in laterality CHD genes is higher in those with laterality CHD compared to non-laterality CHD.

To evaluate whether the identified digenic combinations were specific to the laterality CHD cohorts that we studied, we used GS data from 602 healthy trios (1,806 individuals) available from the 1000G as a control cohort.^21^ We applied the same variant filtering approach and the same strategy for the trans-heterozygous selection of digenic variants. The proportion of families and/or individuals presenting trans-heterozygous digenic variants were then compared between the laterality CHD cohorts and the control cohort. Additionally, the proportion of cases with non- laterality CHD with digenic variants from the Kids First and PCGC cohorts were compared to the proportions with laterality CHD. P-values were calculated using the two-sided Fisher’s exact test (*fisher.test* function in R).

### Biological functional annotation in all mutated genes in the three cohorts

To determine significantly enriched biological processes and pathways, biological functional annotation was performed by Enrichr web-server (http://amp.pharm.mssm.edu/Enrichr).^22^ The most significantly enriched gene ontology (GO) biological process terms were identified in all mutated genes as evidenced by their associated *p*-value.

### Evaluation of identified digenic pairs using digenic predictor (DiGePred)

The digenic prediction score for each digenic pair was calculated by the digenic pair prediction tool (DiGePred), a high-throughput machine learning approach for evaluating the likelihood that dysfunction of gene pairs leads to disease phenotypes. A detailed description of the methods used for digenic prediction has been previously described.^23^ Briefly, DiGePred predicts a digenic score for each gene pair using six network and functional features such as, pathway similarity, co-expression rank across multiple tissue and “network distances” between the genes on protein-protein, pathway, and literature-mined interaction networks from the UCSC gene and pathway interaction browser database. Digenic score threshold was determined for the DiGePred classifier for classifying gene pairs as digenic. The most confident DiGePred threshold determined was F_0.5_ metric which is maximized at a digenic score of 0.496. A gene pair was deemed a candidate digenic pair if the digenic score met the F_0.5_ threshold, i.e., digenic pair was calculated as having the score of >= 0.496. In this analysis, all potential gene pairs available from the 115 genes were used as a background set of genes.

### Cardiac phenotype classification

All cardiac phenotypes were classified by a pediatric cardiologist (S.A.M.) based on available data. Primary CHD lesions were separated into general groups, then laterality CHD was classified into subgroups. Laterality defects included all lesions with malposition or transposition of the great arteries, abnormal ventricular looping, or situs abnormalities of the viscera, thorax, or atria. Among those not classified as laterality CHD, groups were: right-sided lesions, left-sided lesions, conotruncal defects, endocardial cushions defects, Epstein anomaly, and other. Classifications were mutually exclusive, and hierarchies were based upon the Core Cardiac Lesion Outcome Classifications system (C-CLOC).^24^ Within the laterality CHD subjects, subphenotypes were: D-transposition of the great arteries (DTGA), double outlet right ventricle (DORV) with malposed great arteries, congenitally corrected TGA (CCTGA), anatomically corrected TGA, double inlet left ventricle (DILV), tricuspid atresia with malposed great arteries, mitral atresia with malposed great arteries, other L-looped ventricles, situs inversus with CHD, heterotaxy with right atrial isomerism, and heterotaxy with left atrial isomerism.

### Computational and statistical analyses

R and R Studio were used for most computational analyses. Frequency comparisons were performed using the two-sided Fisher’s exact test (*fisher.test* function in R). A post-hoc analysis comparing distribution of digenic variants by race/ethnicity among cohort was performed to determine if the variance in digenic proportions between cohorts could be partially explained by different racial and ethnic distributions between cohorts.

## Results

### Digenic variants identified in three laterality defect cohorts

In the BCM-GREGoR cohort, we performed proband-only exome analysis in 247 unsolved cases with laterality defects and complex CHD followed by Sanger sequencing for variant allele confirmation and segregation analysis in available family members. Variant allele confirmation and segregation analysis indicated likely trans-heterozygous digenic variants in 7 probands (**Table 1**, **Figure 2B, and Figure S1**). The parents of these 7 probands were unaffected. In another extended family of 8 members, the female proband was found to have *DNAAF2/DNAH5* digenic variants, with both alleles inherited from the affected one parent. Of note, variants were not found together in other unaffected 6 family members which included 3 siblings, and unaffected one parent of affected individual, and unaffected maternal grandparents (**Figure S1)**. Therefore, digenic variants were present in 8/247 probands (3.2%). Detailed phenotypes of all the probands with digenic variants in the BCM-GREGoR cohort are described in **Table S3.**

**Figure 2:**
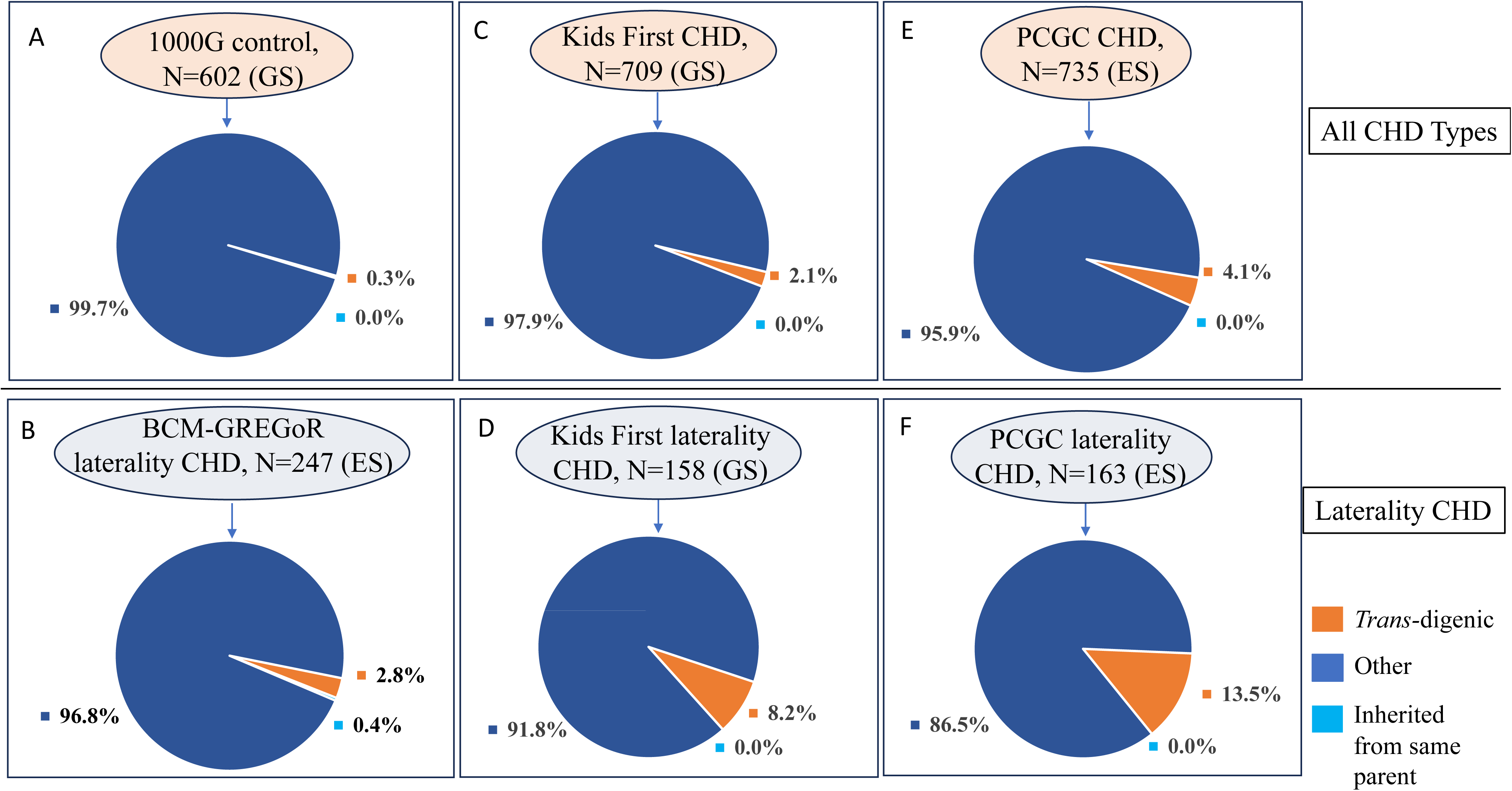
Percentage of trans-heterozygous digenic variants identified in different cohorts. Trans-heterozygous digenic variants were identified in **(A)** 0.3% of control population (2 of 602 trio GS data from the 1000 Genomes Project), (B) 2.8% (7/247) of laterality CHD cases in the BCM-GREGoR cohort, and 0.4% (1/247) of laterality CHD cases in the BCM-GREGoR cohort inherited both variants from affected parent **(C)** 2.1% (of cases in all CHD types in Kids First, and **(D)** 8.2% of the laterality CHD cases in the Kids First cohort, **(E)** 4.1% cases in all CHD types, and **(F)** 13.5% of the laterality cases in the PCGC cohort.

**Table 1:** Digenic variants in BCM-GREGoR cohort.

| ID | Sex | Race/<br>Ethn | Phenotype (detailed) | Atrial<br>situs | Ventricular<br>situs | Arterial<br>situs | CHD<br>group | Gene | Genomic Position | Zyg | C change | P change | Mut | vR | tR | CADD<br>phred | gnomAD | pLI | Clin<br>Var | DiGe<br>Pred<br>score | Disease classified in<br>OMIM |
| --- | --- | --- | --- | --- | --- | --- | --- | --- | --- | --- | --- | --- | --- | --- | --- | --- | --- | --- | --- | --- | --- |
| LAT1663 | F | NHB | TA, DORV, L-MGA | S | D | L | Complex<br>Tri Atresia | *DNAH1* | 3:52365245; T>G | Het | c.T953G | p.L318R | ms | 70 | 145 | 26 | 7.28E-05 | 0 | US | 0.66 | ? Primary ciliary dyskinesia 37 |
|  |  |  |  |  |  |  |  | *DNAH6* | 2:84864340; A>G | Het | c.A4660G | p.R1554G | ms | 39 | 95 | 26 | 2.17E-05 | 0 | US |  | NA |
| LAT1357 | M | H | Unk. | Unk. | Unk. | Unk. | Unk. | *DNAH9* | 17:11713593; G>A | Het | c.G8614A | p.G2872R | ms | 87 | 163 | 32 | 4.61E-05 | 0 | NF | 0.71 | Primary ciliary dyskinesia 40 |
|  |  |  |  |  |  |  |  | *TTC12* | 11:113220878; T>G | Het | c.T1238G | p.L413R | ms | 100 | 193 | 22 | 1.77E-05 | 0 | NF |  | Primary ciliary dyskinesia 45 |
| LAT1085 | F | H | HTX, LAI, L-ventricular looping, CAVC, DORV, L-MGA | A | L | I | HTX (LAI) | *DNAH6* | 2:84881906; A>C | Het | c.A5757C | p.K1919N | ms | 63 | 123 | 15 | . | 0 | NF | 0.53 | NA |
|  |  |  |  |  |  |  |  | *RSPH4A* | 6:116953588; A>G | Het | c.A2135G | p.E712G | ms | 30 | 57 | 23 | 0.0001 | 0 | CI |  | Primary ciliary dyskinesia 11 |
| LAT1031 | F | H | DTGA | S | D | D | DTGA | *DNAH1* | 3:52426708; C>G | Het | c.10278+3C>G | - | sp | 51 | 92 | - | 3.82E-05 | 0 | CI | 0.56 | ? Primary ciliary dyskinesia 37 |
|  |  |  |  |  |  |  |  | *DNAH11* | 7:21765456; C>T | Het | c.C7294T | p.R2432W | ms | 35 | 5 | 25 | 0.0004 | 0 | CI |  | Primary ciliary dyskinesia 7 |
| LAT0882 | F | NHW | DORV, D-MGA, PS | S | D | D | DORV + MGA | *DRC1* | 2:26644243; G>A | Het | c.G331A | p.E111K | ms | 9 | 31 | 25 | 4.37E-05 | 0 | NF | 0.72 | Primary ciliary dyskinesia, 21 |
|  |  |  |  |  |  |  |  | *GAS8* | 16:90106726; G>A | Het | c.G1030A | p.E344K | ms | 13 | 28 | 21 | 4.31E-05 | 0 | US |  | Primary ciliary dyskinesia, 33 |
| LAT0279 | F | H | L-ventricular looping, DILV, LTGA, PS, SubPS | S | L | L | DILV | *DNAH6* | 2:84822780; A>T | Het | c.A2735T | p.D912V | ms | 25 | 44 | 23 | . | 0 | NF | 0.54 | NA |
|  |  |  |  |  |  |  |  | *DNAH9* | 17:11622703; G>A | Het | c.G5605A | p.A1869T | ms | 24 | 52 | 26 | 6.77E-05 | 0 | US |  | Primary ciliary dyskinesia, 40 |
| LAT0230<br>(Daughter of LAT0227) | F | NHW | CAVC, D-MGA, PA | Unk. | Unk. | Unk. | Unk. | *DNAAF2* | 14:50094798; A>G | Het | c.A1939G | p.M647V | ms | 61 | 143 | 16 | . | 0 | US | 0.61 | Primary ciliary dyskinesia, 10 |
|  |  |  |  |  |  |  |  | *DNAH5* | 5:13708372-13708375; del | Het | c.13194-13197del | p.4398_4399del | fs del | 21 | 57 | - | 2.44E-05 | 0 | P/L P |  | Primary ciliary dyskinesia, 3 |
| LAT0227<br>(Parent of LAT0230) | M | NHW | Unk. | Unk. | Unk. | Unk. | HTX | *DNAAF2* | 14:50094798; A>G | Het | c.A1939G | p.M647V | ms | 61 | 143 | 16 | . | 0 | US | 0.61 | Primary ciliary dyskinesia, 10 |
|  |  |  |  |  |  |  |  | *DNAH5* | 5:13708372-13708375; del | Het | c.13194-13197del | p.4398_4399del | fs del | 21 | 57 | - | 2.44E-05 | 0 | P/L P |  | Primary ciliary dyskinesia, 3 |
| LAT0015 | M | H | HTX, RAI, unbalanced CAVC, DORV with D-MGA, PS, SubPS | A | D | D | HTX (LAI) | *CCDC40* | 17:78055582; C>G | Het | c.C1800G | p.D600E | ms | 23 | 58 | 16 | 3.22E-05 | 0 | US | 0.72 | Primary ciliary dyskinesia, 15 |
|  |  |  |  |  |  |  |  | *DNAH1* | 3:52388952; G>A | Het | c.G3574A | p.E1192K | ms | 103 | 198 | 28 | 3.92E-05 | 0 | US |  | Primary ciliary dyskinesia, 37 |

**Abbreviations:** A: Situs Ambiguous; CAVC: Common atrioventricular septal defect; CCTGA: Congenitally corrected transposition of the great arteries; CHD: Congenital heart defects; CI: Conflicting interpretation of pathogenicity; D-MGA: Dextro-malposed great arteries; DILV: Double inlet left ventricle; DORV: Double outlet right ventricle; DTGA: Dextro-transposition of the great arteries; Ethn: Ethnicity; F: Female; H: Hispanic; Het: heterozygous; HTX: Heterotaxy; LAI: Left atrial isomerism; L-MGA: Levo-malposed great arteries; LP: Likely pathogenic; LTGA: Left transposition of the great arteries; M: Male; ms: Missense; Mut: Mutation type; NA: Not available; NF: Not found; NHB: Non-hispanic black; NHW: Non-hispanic white; OMIM: Online mendelian inheritance in men; P: Pathogenic; PS: Pulmonary stenosis; PA: Pulmonary artery Atresia; RAI: Right atrial isomerism; S: Situs Solitus; SubPS: Subpulvular pulmonary stenosis; TA: Tricuspid atresia; tR: Total read; vR: Variant read; Unk.: Unknown; US: Unknown significance; VSD: Ventricular Septal Defect; Zyg: zygosity

In the Kids First and PCGC laterality CHD cohorts, trans-heterozygous digenic variants were identified in 13 (8.2%) and 22 (13.5%) probands, respectively (**Tables 2 and 3**, **Figure 2D and 2F**). In contrast, in the Kids First and PCGC non-laterality CHD cohorts, trans-heterozygous digenic variants were identified in 2 (0.4%) and 8 (1.4%) cases, respectively (**Table S1**). Therefore, in all the CHD cases studied, digenic variants were identified in 2.1% and 4.1% cases available from the Kids First and PCGC datasets, respectively (**Figure 2C and 2E**). In these 3 cohorts, 106 SNVs (101 missense, 1 splice site, and 4 frameshift) were detected in 54 cases (53 families) (**Figure S2).**

**Table 2:** Digenic variants in Kids First cohort.

| **Digenic variants in laterality CHD** |  |  |  |  |  |  |  |  |  |  |  |  |  |  |  |  |  |  |  |  |  |
| --- | --- | --- | --- | --- | --- | --- | --- | --- | --- | --- | --- | --- | --- | --- | --- | --- | --- | --- | --- | --- | --- |
| ID | Sex | Race/<br>Ethn | Phenotype<br>(detailed) | Atrial<br>situs | Ventricular<br>situs | Arterial<br>situs | CHD<br>group | Gene | Genomic Position | Zyg | C change | P change | Mut | vR | tR | CADD<br>phred | gnomA<br>D | pLI | Clin<br>Var | DiGe<br>Pred<br>score | Disease classified in<br>OMIM |
| BS_7JBDM67P | M | NHW | DTGA, DPVC | S | D | D | DTGA | *DNAH5* | 5:13811745; G>A | Het | c.C7309T | p.R2437C | ms | 9 | 22 | 24.7 | 4.46E-05 | 0 | CI | 0.61 | Primary ciliary dyskinesia 3 |
|  |  |  |  |  |  |  |  | *DNAH9* | 17:11744983; G>A | Het | c.G6298A | p.A2100T | ms | 9 | 23 | 15.9 | 0.0002 | 0 | US |  | Primary ciliary dyskinesia 40 |
| BS_SFBX37TD | M | HW | DTGA, PS | S | D | D | DTGA | *DNAH9* | 17:11647103; C>T | Het | c.C2002T | p.R668W | ms | 23 | 42 | 29.8 | 2.48E-05 | 0 | NF | 0.57 | Primary ciliary dyskinesia 40 |
|  |  |  |  |  |  |  |  | *DNAH11* | 7:21748740; C>T | Het | c.C8671T | p.R2891W | ms | 19 | 40 | 35 | 0.0002 | 0 | US |  | Primary ciliary dyskinesia 7 |
| BS_ST7TSHW0 | F | HW | DTGA, PS, LSVC connecting to the CS | S | D | D | DTGA | *MEGF8* | 19:42359134; G>T | Het | c.G5380T | p.G1794W | ms | 11 | 23 | 23.3 | 3.98E-05 | 0 | NF | 0.004 | Carpenter syndrome 2 |
|  |  |  |  |  |  |  |  | *DNAH1* | 3:52390959; C>G | Het | c.C9646G | p.L3216V | ms | 19 | 27 | 23.5 | 0.001 | 0 | US |  | ? Primary ciliary dyskinesia 37 |
| BS_998BPYWT | F | NHA | DTGA, right aortic arch with mirror image branching | S | D | D | DTGA | *SHROOOM3* | 4:76739414; G>A | Het | c.G1241A | p.R414Q | ms | 8 | 29 | 24.8 | 0.0002 | 0 | NF | 0.51 | NA |
|  |  |  |  |  |  |  |  | *DNAH9* | 17:11617516; C>T | Het | c.C1010T | p.P337L | ms | 14 | 3 | 23.7 | 0.0002 | 0 | CI |  | Primary ciliary dyskinesia 40 |
| BS_ABW87ZXQ | F | HW | DTGA, PS, VSD, LVOT | S | D | D | DTGA | *DNAH9* | 17:11886927; C>T | Het | c.C11074T | p.L3692F | ms | 22 | 44 | 33 | 0.0001 | 0 | LB | 0.001 | Primary ciliary dyskinesia 40 |
|  |  |  |  |  |  |  |  | *NPHP3* | 3:132689196; C>T | Het | c.G2761A | p.E921K | ms | 19 | 34 | 23.1 | . | 0 | NF |  | Nephronophthisis 3, Meckel synd. 7, Renal-hepatic-pancreatic dysplasia 1 |
| BS_VTKKFCRS | F | NHW | DORV, D-MGA, CA, LSVC non communicating with RSVC, HLV | S | D | D | DORV w/ D-MGA | *DNAH5* | 5:13776582; C>A | Het | c.G9230T | p.R3077L | ms | 19 | 37 | 34 | 0.0001 | 0 | CI | 0.58 | Primary ciliary dyskinesia 3 |
|  |  |  |  |  |  |  |  | *DNAH6* | 2:84762905; G>A | Het | c.G10663A | p.A3555T | ms | 15 | 38 | 23 | 6.19E-05 | 0 | NF |  | NA |
| BS_84AG8G2W | M | NHW | DORV, D-MGA, MA, VSD, LSVC connecting to the CS, HLV | S | D | D | DORV w/ D-MGA | *KIF7* | 15:89633796; C>T | Het | c.G2482A | p.V828M | ms | 22 | 38 | 32 | 0.0002 | 0 | US | 0.0001 | Joubert syndrome 12 |
|  |  |  |  |  |  |  |  | *RSPH4A* | 6:116617207; C>G | Het | c.C584G | p.P195R | ms | 22 | 45 | 18.6 | 0.0006 | 0 | US |  | Primary ciliary dyskinesia 11 |
| BS_FAY6B86G | F | HB | DORV, PA, MA, HLV, right aortic arch with mirror image branching | S | D | D | DORV (likely MGA) | *DNAH9* | 17:11969430; A>G | Het | c.A13364G | p.Q4455R | ms | 22 | 34 | 15.3 | 7.81E-05 | 0 | US | 0.52 | Primary ciliary dyskinesia 40 |
|  |  |  |  |  |  |  |  | *PKD1L1* | 7:47905323; C>T | Het | c.G1525A | p.V509I | ms | 19 | 42 | 15.1 | 6.88E-05 | 0 | LB |  | Heterotaxy, visceral, 8 |
| BS_6WZV64SA | F | NHW | TA, VS?, DTGA, CoA, L-loop ventricles, DC | S | L | D | Complex Tri Atrisia | *CCDC39* | 3:180659549; C>T | Het | c.G641A | p.R214H | ms | 18 | 35 | 26.6 | 9.73E-05 | 0 | US | 0.82 | Primary ciliary dyskinesia 14 |
|  |  |  |  |  |  |  |  | *CCNO* | 5:55233337; A>G | Het | c.T187C | p.S63P | ms | 9 | 21 | 24 | 0.0007 | 0 | US |  | Primary ciliary dyskinesia 29 |
| BS_BBV8D7ST | M | NHW | LTGA, VSD | S | L | L | CCTGA | *MEGF8* | 19:42348314; G>A | Het | c.G2140A | p.A714T | ms | 13 | 25 | 16.6 | 0.0003 | 1 | NF | 0.52 | Carpenter syndrome 2 |
|  |  |  |  |  |  |  |  | *KIF7* | 15:89632900; G>A | Het | c.C2815T | p.R939W | ms | 19 | 36 | 34 | 7.71E-05 | 0 | US |  | Joubert syndrome 12 |
| BS_CNA0GMS5 | F | NHW | HTX, DORV, L-MGA, PS, DC, asplenia | A | L | L | HTX (RAI) | *FLNA* | X:154360184; G>A | Het | c.C3611T | p.P1204L | ms | 18 | 38 | 22 | 1.41-05 | 0.99 | CI | 0.02 | Cardiac valvular dysplasia, X-linked |
|  |  |  |  |  |  |  |  | *HYDIN* | 16:70872081; G>A | Het | c.C11047T | p.R3683W | ms | 10 | 22 | 30 | 0.001 | NA | US |  | Primary ciliary dyskinesia 5 |
| BS_0R2CQ47C | M | NH* | HTX, DORV, D-MGA, ASD, MA, VSD, HLV | A | D | D | HTX (? type) | *CFAP46* | 10:132847067; C>A | Het | c.G6132T | p.E2044D | ms | 20 | 35 | 24 | 2.48E-06 | 0 | NF | 0.62 | NA |
|  |  |  |  |  |  |  |  | *DNAH8* | 6: 38875744; C>T | Het | c.C7774T | p.R2592W | ms | 10 | 24 | 17.7 | 0.001 | 0 | US |  | Spermatogenic failure 46 |
| BS_8E1NXAPG | M | NHB | HTX, single RV, DIRV, hypoplastic MV | Unk. | Unk. | Unk. | HTX (? type) | MRPS25 | 3:15052554; G>A | Het | c.C409T | p.R137W | ms | 19 | 37 | 35 | 0.0002 | 0 | NF | 0.54 | ?Combined oxidative phosphorylation deficiency 50 |
|  |  |  |  |  |  |  |  | DNAH6 | 2:84642024; A>G | Het | c.A5048G | p.K1683R | ms | 15 | 38 | 27.2 | 1.42E-05 | 0 | NF |  | NA |

| Digenic variants in non-laterality CHD |  |  |  |  |  |  |  |  |  |  |  |  |  |  |  |  |  |  |  |  |  |
| --- | --- | --- | --- | --- | --- | --- | --- | --- | --- | --- | --- | --- | --- | --- | --- | --- | --- | --- | --- | --- | --- |
| ID | Sex | Race/ Ethn | Phenotype (detailed) | Atrial situs | Ventricular situs | Arterial situs | CHD group | Gene | Genomic Position | Zyg | C change | P change | Mut | vR | tR | CADD phred | gnomAD | pLI | Clin Var | DiGe Pred score | Disease classified in OMIM |
| BS_73YTS16K | M | NHW | VSD, TOF, PA, PFO, Hypoplastic left pulmonary artery, PS | S | D | S | CONO | DNAH5 | 5:13923336; C>T | Het | c.G382A | p.V128M | ms | 23 | 38 | 23.6 | 0.0003 | 0 | CI | 0.54 | Primary ciliary dyskinesia 3 |
|  |  |  |  |  |  |  |  | DNAAF4 | 15:55467113; G>A | Het | c.C454T | p.R152W | ms | 11 | 26 | 34 | 0.0003 | NA | US |  | Primary ciliary dyskinesia 25 |
| BS_CGZN5KMX | M | HW | Bicuspid aortic valve, aortic stenosis and regurgitation mitral valve stenosis | S | D | S | LSL | DNAH11 | 7:21750312; C>A | Het | c.C8888A | p.S2963Y | ms | 20 | 46 | 28.3 | 0.0005 | 0 | CI | 0.57 | Primary ciliary dyskinesia 7 |
|  |  |  |  |  |  |  |  | DNAH9 | 17:11632630; A>T | Het | c.A1562T | p.D521V | ms | 18 | 37 | 24 | 3.35E-05 | 0 | US |  | Primary ciliary dyskinesia 40 |

**Table 3:**
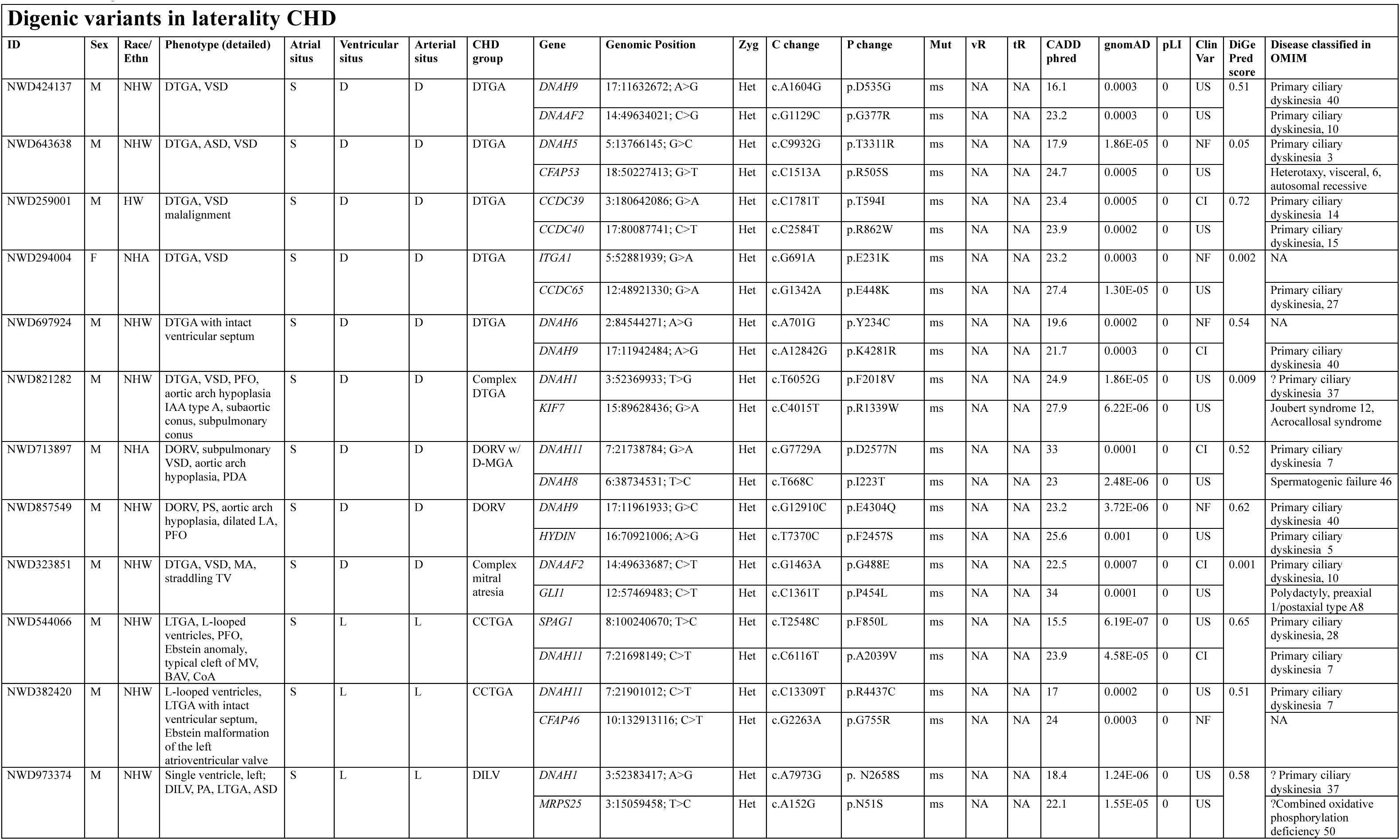
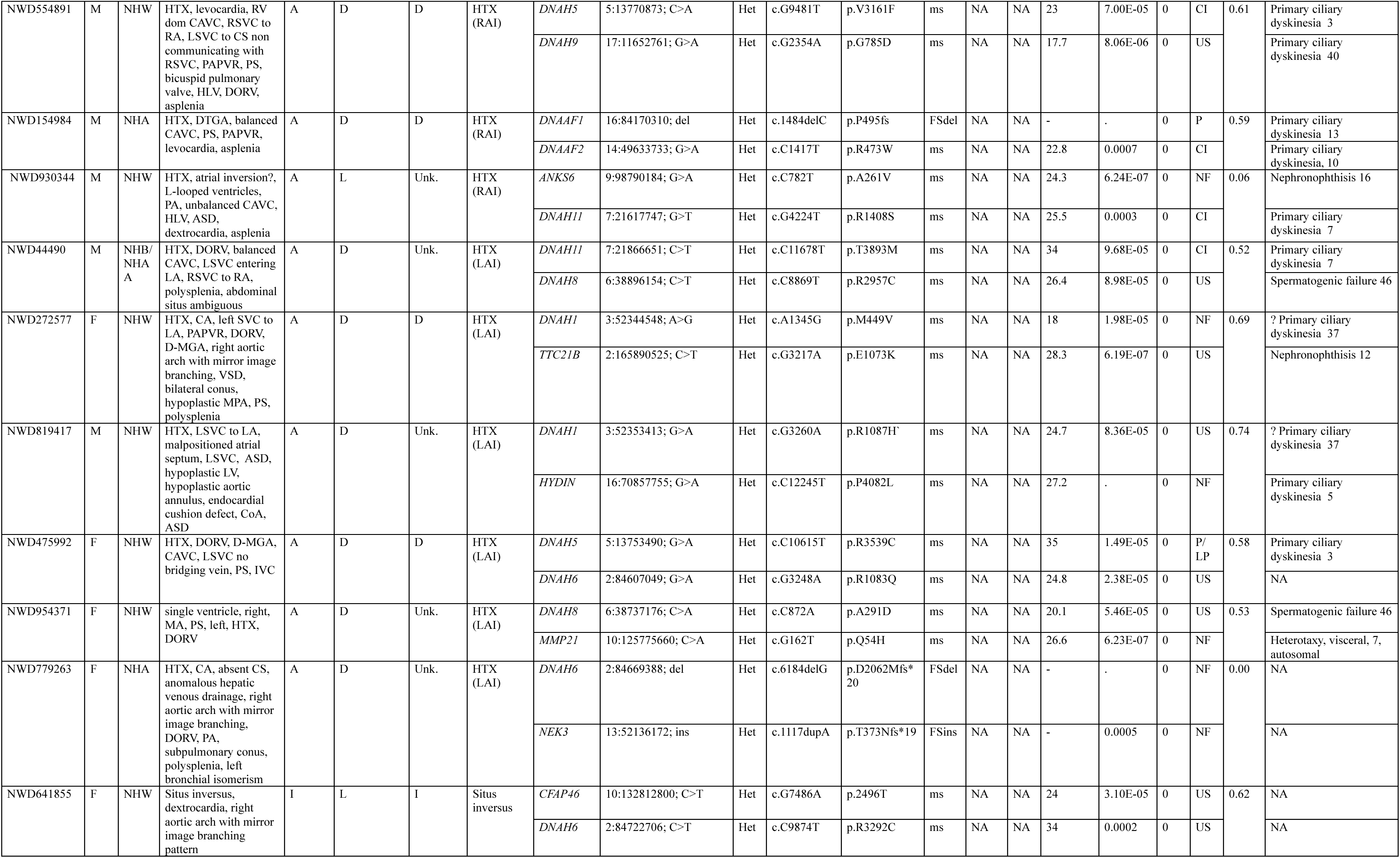
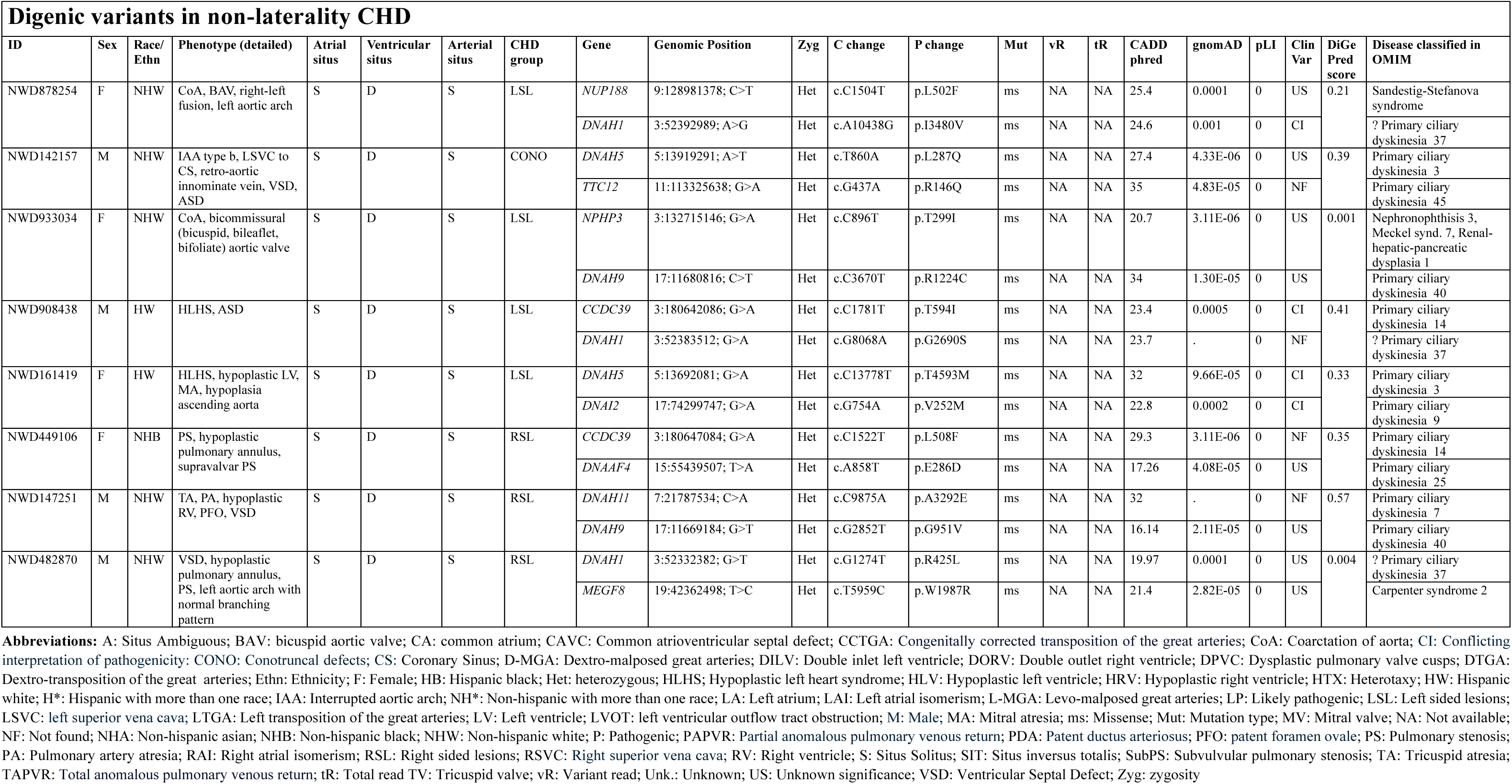
Digenic variants in PCGC cohort.

Biological functional annotation of the 36 mutated genes identified in these three cohorts revealed that they were most significantly enriched in cilium movement (*p =* 0.0003), axoneme assembly (*p =* 0.0007), cilium-dependent cell motility (*p* = 0.0021), cilium assembly (*p* = 0.0057), axonemal dynein complex assembly (*p* = 0.0079), and cellular component assembly (*p* = 0.0096), indicating that all are enriched in ciliary body pathway (**Figure 3**).

**Figure 3:**
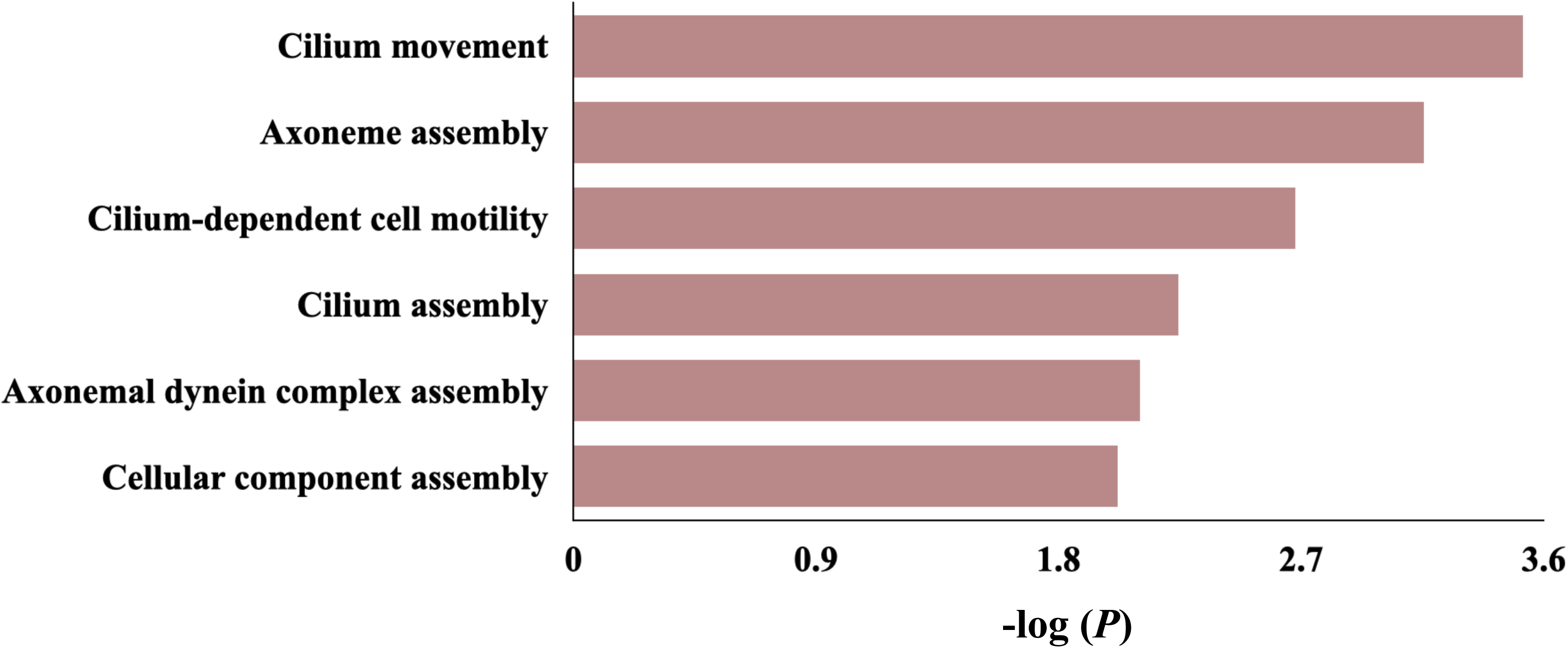
Biological functional annotation of the identified genes with trans-heterozygous digenic variants in three laterality cohorts. Our analysis revealed that they were most significantly enriched in cilium movement, axoneme assembly, cilium-dependent cell motility, cilium assembly, axoneme dynein complex assembly, and cellular component assembly with their –log (*P*) shown on the x-axis.

### Prevalence comparisons between laterality CHD, control cohort, and non-laterality CHD cohorts

Among the 602 cases from the healthy population in the 1000G (control cohort), 2 individuals were found to have variant combinations satisfying the criteria we established for the digenic events (gene, variant and parental inheritance), for a prevalence of 0.3%. This proportion was significantly less than that observed in the laterality CHD cohorts (BCM-GREGoR, 3.2%; Kids First, 8.2%; PCGC, 13.5*%; p* = 0.001, 1.4e-07, and 8.9e-13 respectively).

In the non-laterality CHD cohorts, prevalence of qualifying digenic variants was 0.4% (Kids First) and 1.4% (PCGC). This was not significantly different from observed in controls *(p* = 1 and 0.059 respectively, Fisher’s exact test), but was significantly lower than the frequency observed in the laterality CHD cohorts (Kids First non-laterality CHD 0.4% versus laterality CHD 8.2%, *p =* 3.3e- 07; PCGC non-laterality CHD 1.4% versus laterality CHD 13.5%, *p* = 8.9e-09).

These comparisons show significantly higher prevalence of combined rare deleterious variants in the known laterality defect genes in the laterality CHD cohorts than in either control or non- laterality CHD cohorts, providing further evidence that digenic inheritance is a clinically relevant model in laterality CHD.

### Implication of ciliary genes in pathogenesis of laterality defect

Out of 55 (48%) ciliary, and 60 (52%) non-ciliary genes considered for analysis of laterality defect cases using digenic inheritance approach in this study, 27 (49%), and 9 (15%) genes were mutated in those three cohorts *(p* = 0.002), respectively (**Figure 4**, **Figure 5**). Also, 74% (20/27) of mutated ciliary genes encode ciliary motility related proteins. In contrast, 15% (4/27), 4% (1/27), and 7% (2/27) of mutated ciliary genes encode ciliary nonmotility related proteins, ciliogenesis, and trafficking related proteins (proteins involved in the movement of proteins to and within the primary cilium), respectively. In those three cohorts, there were 43 (7.6%) cases with laterality CHD in total presenting with digenic variants. Remarkably, 33 (77%) out of 43 cases had both digenic candidate variants in genes related to the cilia. The majority of variants were found in genes encoding for ciliary proteins, of which most encode for components of the ciliary motility apparatus such as outer dynein arm (*DNAH1, DNAH5, DNAH8, DNAH9, DNAH11*, and *SPAG1*), inner dynein arm (*DNAH6*), nexin-dynein regulatory complex (*GAS8, DRC1*, *CCDC39*, and *CCDC40*), radial spoke (*RSPH4A*), and central pair (*HYDIN*). Out of 33 ciliary digenic pairs identified in laterality cases, 23 correspond to the ciliary motility apparatus, of which 10 ciliary digenic pairs were in the same structural complex of motile cilia, including *DNAH1/DNAH11, DNAH9/DNAH11, DNAH5/DNAH9 (*in two cases)*, SPAG1/DNAH11, DNAH8/DNAH11* (in two cases) (outer dynein arm), *DRC1/GAS8, CCDC39/CCDC40* (nexin–dynein regulatory complex), and *DNAAF1*/*DNAAF2* (dynein pre-assembly complex) (**Figure 5**).

**Figure 4:**
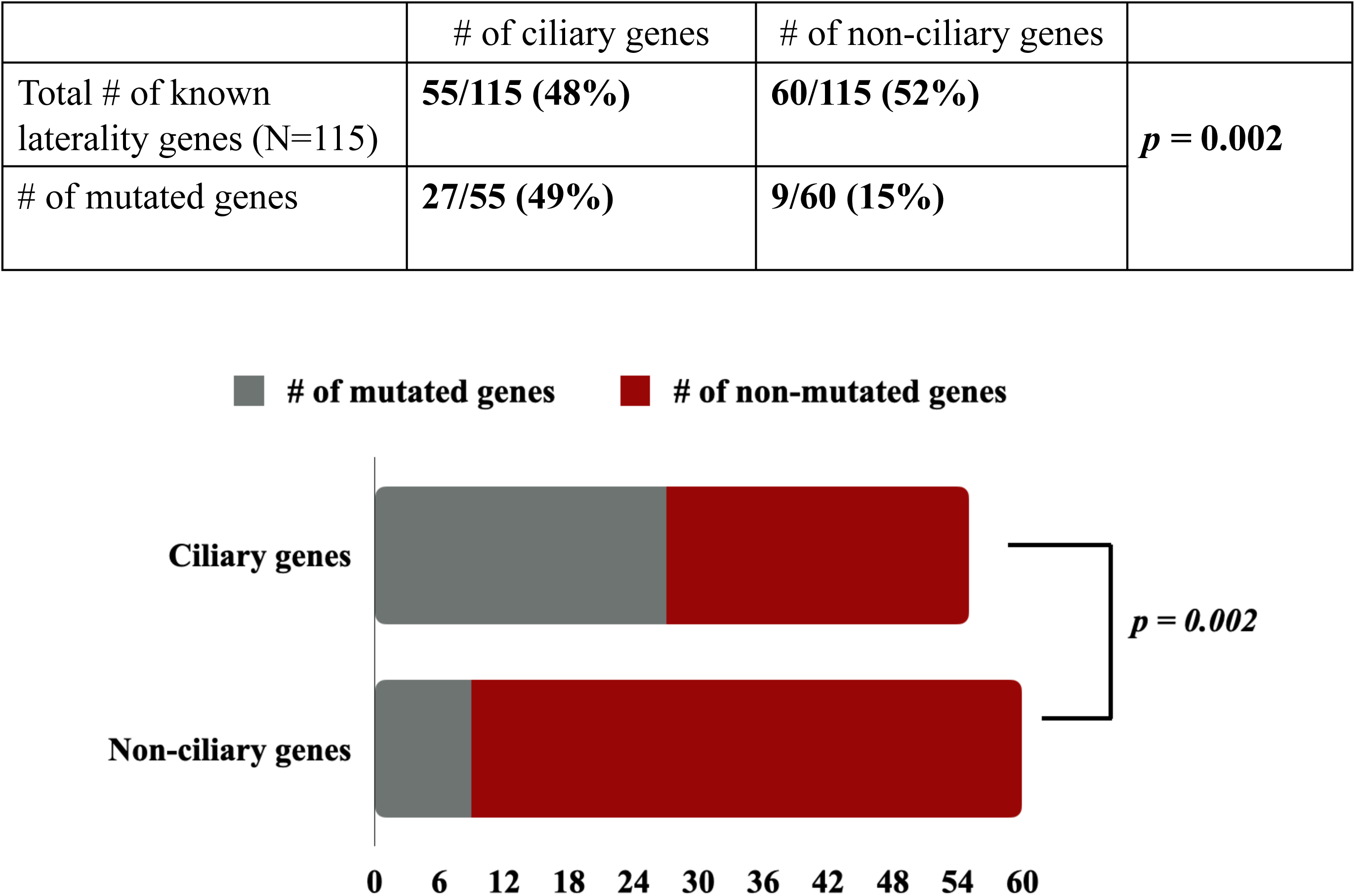
Proportion of trans-heterozygous digenic variants in ciliary genes as compared to non-ciliary genes. In three laterality cohorts, the proportion of digenic variants present in ciliary genes (27/55) is significantly higher than non-ciliary genes (9/60), as indicated by *p =* 0.002.

**Figure 5:**
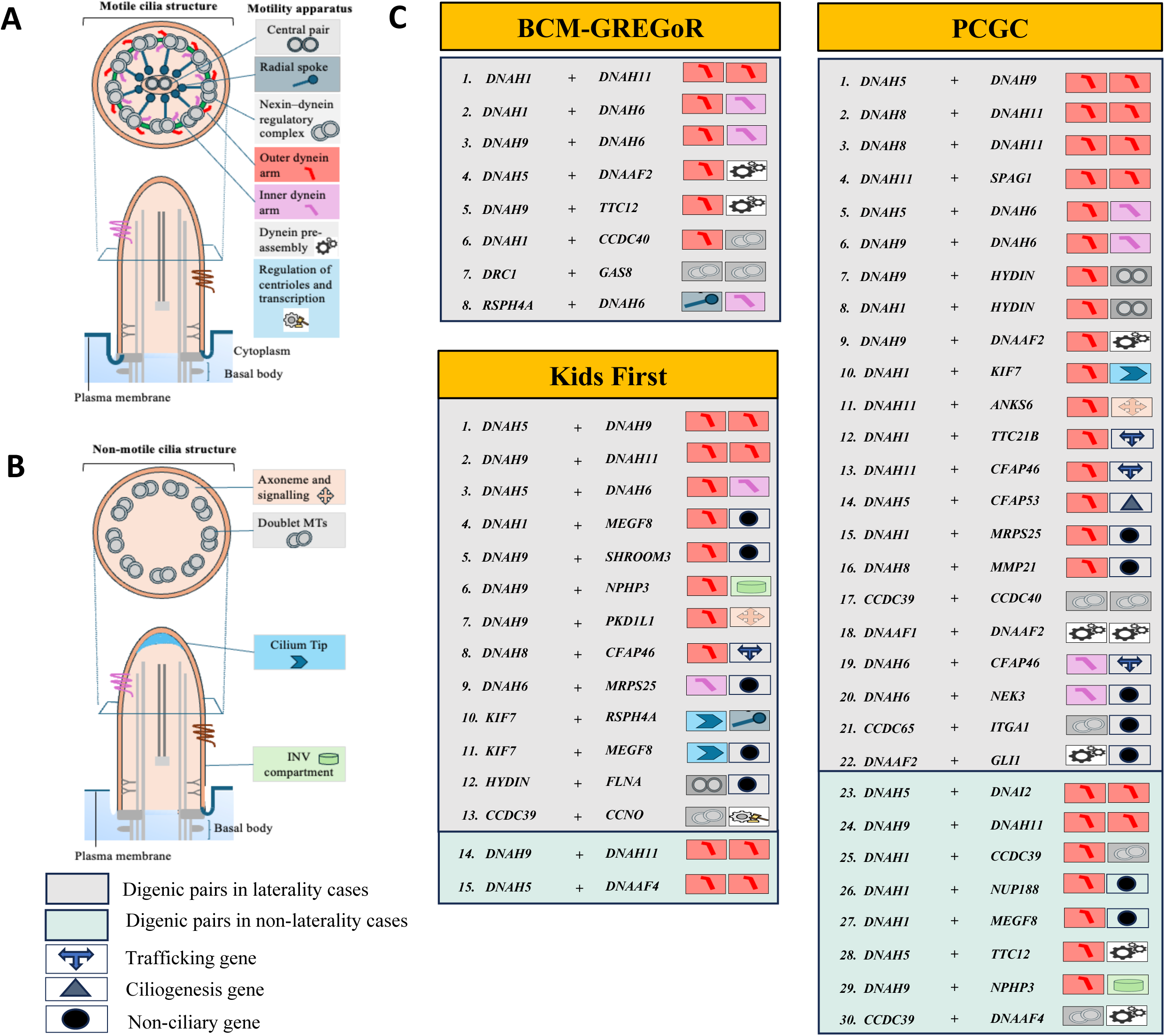
Structural and functional features of identified ciliary and non-ciliary genes in probands. **(A)** Structure of motile cilia, **(B)** structure of non-motile cilia, **(C)** gene pairs in gray boxes are representing those, which were identified to carry trans-heterozygous digenic variants in laterality cases in three cohorts, gene pairs in green boxes are representing those, which were identified to carry trans-heterozygous digenic variants in non-laterality cases in three cohorts.

### Frequency analysis by laterality CHD subtype and race/ethnicity

The three laterality CHD cohort had highly variable distributions of laterality CHD subtypes (**Table S4**). Digenic variants were most common in mitral atresia with malposed great arteries (3/6, 50%), heterotaxy with right atrial isomerism (7/76, 9.2%), and heterotaxy with left atrial isomerism (6/53, 11.3%). The variation in the prevalence of digenic variants between the cohorts did not seem to be explained by variance in laterality CHD subtypes. For example, even though the proportion of probands with isomerism was fairly similar (28.3%, 18.4%, and 18.4% for BCM-GREGoR, Kids First, and PCGC respectively), the digenic variant prevalence among these varied greatly (5.7%, 3.4%, 26.7% respectively).

When examining the cohorts by race/ethnicity, the cohorts were significantly different (*p*<0.0001), with the BCM-GREGoR cohort having a larger proportion of Hispanic probands (34.8% compared to 19.6% and 14.7% respectively in Kids First and PCGC respectively, **Table S5**). However, there was no significant difference in proportion of digenic variants by race/ethnicity (*p* = 0.096). While Non-Hispanic Asian subjects had the highest proportion of digenic variants (22.7%), this was mainly driven by the PCGC cohort, in which 4 of the 8 Asian subjects had digenic variants, while the BCM cohort had 0/7 (0%) and the Kids First cohort had 1/7 (14.3%).

### Evaluation of identified digenic pairs in three laterality defect cohorts

The scores for all digenic pairs were downloaded on February 2024 after submitting our 115 gene list to DiGePred Server (http://servers.meilerlab.org/index.php/servers/show?s_id=28). For the analysis of CHD cohorts, we used “all digenic vs unaffected score”. Of the 39 unique gene pairs identified in three laterality cohorts, 29 gene pairs had digenic score that met the F_0.5_ threshold (digenic score equal to 0.496, **Tables 1-3**). In contrast, only 218 gene pairs out of 6,555 (all potential gene pairs available from the 115 genes) had digenic score >= 0.496 (Fisher exact test, *p* = 2.2e-16) (**Figure 6**). Variants in these 29 digenic pairs were likely to be potential disease causing based on six network and functional features such as, pathway similarity, phenotype similarity, co-expression rank across multiple tissue, and “network distances” between the genes on protein-protein, pathway, and literature-mined interaction networks from UCSC (https://genome.ucsc.edu/) and pathway interaction browser database.

**Figure 6:**
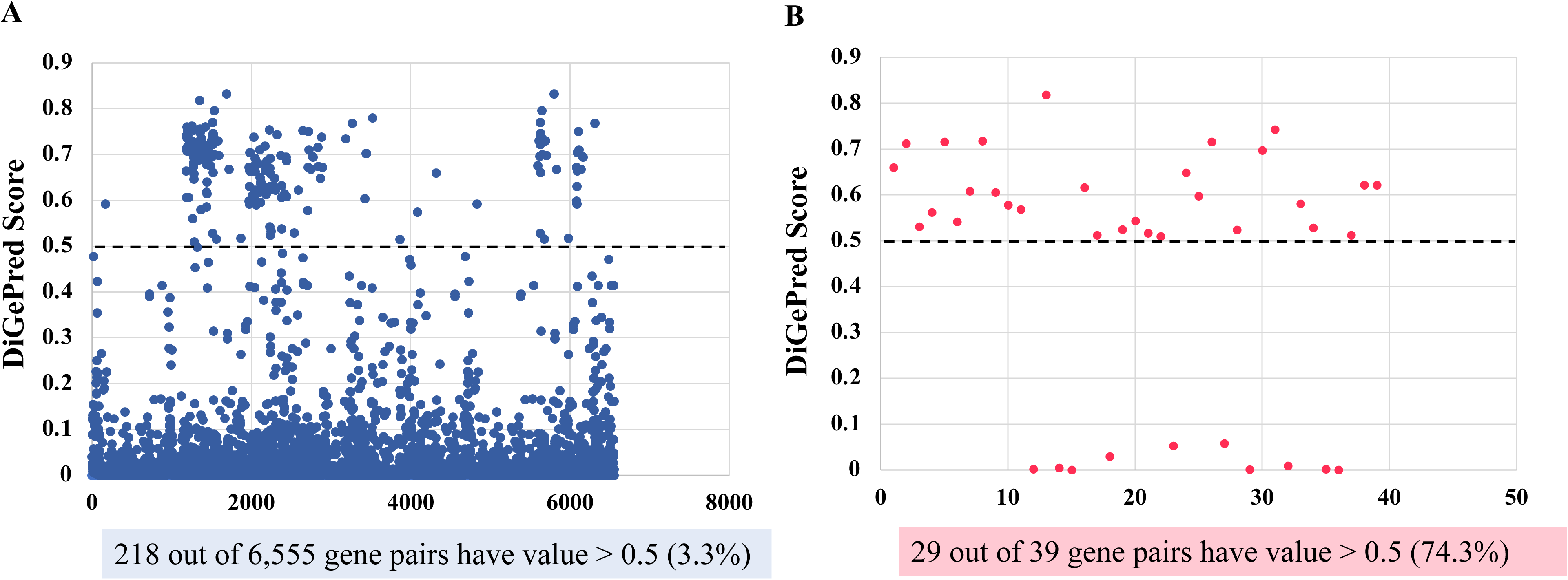
A significantly higher proportion of the genes with the identified trans-heterozygous digenic hits in three cohorts are predicted to be a digenic gene pair. **(A)** Using the previously described threshold (dash lines), 218 (3.3%) out of total 6,555 all potential gene pairs available from the 115 known laterality defect genes are predicted to be digenic hits. **(B**) A significantly higher proportion, 74.3%, 29 out of total 39 unique gene pairs identified in three laterality cohorts are predicted to be digenic hits (*P* = 2.2e-16) compared to the 6,555 all potential gene pairs available from the 115 known laterality defect genes.

When prevalence comparisons were repeated using the stricter digenic criteria, in the BCM-GREGoR cohort, all the identified digenic variants met the F_0.5_ threshold (8/247), which is statistically significant as compared to digenic variants meeting the F_0.5_ threshold in 1000G control cohort (1/602) (*p* = 0.0003). Similarly, statistically significant number of identified digenic variants in Kids First (9/158) and PCGC laterality CHD cohorts (16/163) met the F_0.5_ threshold as compared to digenic variants in 1000G control cohort, *p* = 7.3e-06, and 4.5e-10 respectively. In the non-laterality CHD cohorts, qualifying digenic variants that met the F_0.5_ threshold were 2/551 in Kids First, and 1/572 in PCGC. This was not significantly different from observed in controls *(p* = 0.609 and 1 respectively, Fisher’s exact test).

## Discussion

Human LR asymmetry plays an important role in normal organogenesis and provides the developmental basis for correct heart looping.^25^ LR asymmetry disorders in early embryonic development may result in a series of congenital birth defects including laterality CHD.^26^ Due to genetic and phenotypic heterogeneity, more than 60% of their underlying molecular causes are unknown. In this study, we addressed the relevance of a digenic model for unsolved laterality CHD cases. To delineate the underlying genetic causes of laterality defect, we applied a digenic model approach by screening the personal genome of unsolved cases for variants in 115 known laterality defect genes from three cohorts: BCM-GREGoR (N = 247 proband ES), Kids First (N = 158 trio GS), and PCGC (N = 163 trios ES). Trans-heterozygous digenic variants were observed in 7 (2.8%), 13 (8.2%), and 22 (13.5%) cases with laterality defects in the BCM-GREGoR, Kids First, and PCGC respectively. All the parents of probands with trans-heterozygous digenic variants parents were unaffected. One female proband in BCM-GREGoR carried *DNAAF2*/*DNAH5* digenic variants, both inherited from her one of the affected parent. Notably, these variants were not present together in the six unaffected family members. So, in BCM-GREGoR, there were 7 probands (2.8%) with trans-heterozygous digenic variants, and one proband (0.4%) with inherited digenic variants. Trans-heterozygous digenic variants were identified in 2 (0.4%) and 8 (1.4%) cases in additional 551 and 572 trios with non-laterality CHD in Kids First, and PCGC cohorts respectively.

A higher proportion of digenic variants in known laterality defect genes in laterality defect cases signifying the contribution of digenic variants to laterality defects. All the observed 36 unique mutated genes in three cohorts are known to be inherited in autosomal recessive manner (except *FLNA*), therefore, heterozygous variants in two such genes signifying the digenic inheritance model. Out of 39 unique gene pairs identified in three laterality cohorts, 29 were likely to be potential digenic hits based on their pathway similarity, phenotype similarity, co-expression rank, protein-protein interaction distance, and pathway distance (*p* = 2.2e-16) (**Figure 6**).

Our analysis shows that the proportion of individuals carrying trans-heterozygous digenic variants differs significantly between control cohorts and laterality defect cohorts (*p* = 0.001, 1.4e-07, and 8.9e-13 respectively in BCM-GREGoR, Kids First, and PCGC vs. 1000G). However, no significant difference was observed between controls and cases with non-laterality defects (*p* = 1, 0.059 in Kids First and PCGC vs. 1000G). Furthermore, 48% of ciliary genes were mutated compared to 15% of non-ciliary genes (*p* = 0.002), with 77% of laterality cases with digenic variants have mutated ciliary genes, primarily affecting the ciliary motility apparatus. Ten motile ciliary digenic pairs were in the same structural complex of motile cilia indicating that these gene pairs coding proteins could genetically interact with each other and can affect complex structure and function. Functional interdependence between different types of dyneins have been seen in *Chlamydomonas,* supporting a role for digenic interactions functionally linking outer/inner dynein arm components.^27,28,29^ Out of 27 mutated ciliary genes in three laterality cohorts, 19 were known to be causative for primary ciliary dyskinesis (PCD), a condition that typically causes sinopulmonary disease. This reflects the common requirement for motile cilia in both left-right patterning and airway mucus clearance. We did not have biopsy data or complete clinical data to know if a PCD diagnosis was present in cases. A significant role for ciliary genes was also found in recessive forms of CHD other than laterality defects, suggesting important functions of ciliary genes in cardiac septation^30^. Another possibility is that the lesions classified as non-laterality in the Kids First and PCGC cohort were incompletely phenotyped in the database, and may have underlying unrecognized on unidentified laterality alterations.

Digenic inheritance has been already suggested in laterality defects,^14^ Noonan syndrome,^31^ and hereditary spastic paraplegia.^32^ We observed two cases with digenic variants in *DNAH5*/*DNAH6* gene pairs, one in each Kids First and PCGC laterality cohorts; both cases had phenotypic similarity in terms of DORV and D-malposed great arteries (D-MGA) and were categorized in DORV with complex D-malposition and heterotaxy (left atrial isomerism) CHD groups, respectively. Three cases with heterotaxy have been previously reported with digenic variants in *DNAH5*/*DNAH6* gene pairs, and experimental modeling with double gene knockdown showed digenic interactions of *DNAH6* with *DNAH5* could disrupt motile cilia function in the respiratory epithelia and also cause heterotaxy in zebrafish embryos,^14^ these findings support *DNAH5*/*DNAH6* as a causative gene pair in cases with laterality defects. *DNAH*5 and *DNAH6* encode for components of the ciliary motility apparatus outer dynein arm and inner dynein arm respectively. Also, digenic variants in *DNAH5*/*DNAH9*, *DNAH6*/*DNAH9*, and *DNAH8*/*DNAH11* gene pairs were observed in more than one case in our three cohorts: *DNAH5*/DNAH9; one in each Kids First, and PCGC (DTGA, and heterotaxy with right atrial isomerism, respectively), *DNAH6*/*DNAH9*; one in each BCM-GREGoR, and PCGC (LTGA with DILV, and DTGA, respectively), *DNAH8*/*DNAH11*; two cases in PCGC (DORV with D-MGA, and heterotaxy with left atrial isomerism). Digenic variants in *MEGF8*/*DNAH1*, *DNAH9*/*DNAH11*, and *DNAH9*/*NPHP3* gene pairs were observed in both laterality and non- laterality cases indicating that there may be incomplete penetrance in non-laterality cases or additional phenotypic features should be explored. Overall, the identified digenic variants in three cohort include 106 SNVs (101 missense, 1 non-canonical splice site, and 4 frameshift).

The prevalence of digenic variants was relatively higher in the Kids First and PCGC laterality CHD cohorts as compared to the BCM-GREGoR. The question was raised if this could be due to the difference in racial/ethnic composition among all three cohorts. Among those in the BCM-GREGoR cohort, 34.8% of cases were Hispanic, as compared to 19.6% and 14.7% of the cases in the Kids First and PCGC cohorts respectively. A previous report demonstrated that in the BCM-GREGoR laterality CHD cohort, the most common variant was in the *NODAL* gene (G260R) and 94% of the affected individuals were of Hispanic ethnicity.^33^ However, in this cohort, there was not a significant difference in digenic variants by race/ethnicity.

As limitations of our study, we were not able to obtain access to the structural variants (SV) calls available from the Kids First and PCGC datasets that may further increase the diagnosis rate. Besides this, this study can be extended to the investigation of non-coding variants in any of the studied cohorts.

Collectively, these findings strengthen the hypothesis of digenic inheritance among laterality defects candidate genes in laterality cases and provide further evidence that digenic epistatic interaction can contribute to the complex genetics of laterality defects.

## Data Availability

The datasets used and/or analyzed during the current study are available in the supplementary data.

## Declaration of interests

The authors declare no competing interests.

## Acknowledgements

We would like to thank all the families and patients for their participation as well as the referring genetic counselors and physicians. This work was supported in part by the US National Human Genome Research Institute/National Heart Blood Lung Institute jointly funded Baylor Hopkins Center for Mendelian Genomics (UM1HG006542), by the National Institutes of Health (NIH) (5R01 HD039056, 5R01 HL091771), by the Genomic Research Elucidates the Genetics of Rare disease (GREGoR) program (UM1 HG011758) to J.E.P., J.R.L. and R.A.G., and by the National Institute of Neurological Disorders and Stroke (NINDS R35 NS105078) to J.R.L.. For the Kids First study, dbGaP data with accession <u>phs002517.v4.p2</u> was used. The data from this study phs002517 was made available pre-publication without embargo to support rapid and collaborative research in pediatric cancer via the NCI’s Cancer Research Data Commons (https://datacommons.cancer.gov). This availability is made possible with the support of NCI’s Childhood Cancer Data Initiative (grant No. 3P30CA082103-21S9) and Gabriella Miller Kids First Pediatric Research Program (X01 CA267587). Initial data generation efforts and coordination costs were supported by a number of philanthropic and industry partners with further details at cbtn.org. A.R. was supported by Pediatric Cardiac Genomics Consortium and Cardiovascular Development Data Resource Center 2023 Fellows Program (AGT012018) at the University of Texas at Health Science Center at Houston, TX. Z.C.A was supported by the TOPMed NHLBI fellowship and Simons Foundation pilot award (AGT011737). The content is solely the responsibility of the authors and does not necessarily represent the official views of the NIH.

## Author’s contributions

Conceptualization: A.R., J.R.L., Z.C.A., S.A.M.; Acquisition of data: Z.C.A., J.K., P.J.L., B.D.G., C.E.S., E.B., J.E.P., R.A.G.; Data analysis: A.R., J.Y., K.J.C., J.R.L., Z.D., Z.C.A., S.A.M.; Funding acquisition: Z.C.A., S.A.M., J.R.L., J.E.P., R.A.G.; Visualization: A.R., Z.D., K.J.C.; Clinical data interpretations: S.A.M.; Writing original draft: A.R.; Writing review and editing: all co-authors read, edited and approved the final manuscript.

## Data and materials availability

The datasets used and/or analyzed during the current study are available in the supplementary data.

## Notes

### Competing Interest Statement

The authors have declared no competing interest.

### Author Declarations

Institutional Review Board of Baylor College of Medicine approved research study protocol (IRB approval number: H-1843) and Baylor Genetics (BG) for clinical exomes (IRB approval number: H-48014) for Baylor College of Medicine-Genomics Research to Elucidate the Genetics of Rare Diseases (BCM-GREGoR)data. PCGC subjects were recruited to the Congenital Heart Disease Genetic Network Study of the Pediatric Cardiac Genomics Consortium (CHD GENES: ClinicalTrials.gov identifier NCT01196182). The institutional Review Boards of Boston's Children's Hospital, Brigham and Women's Hospital, Great Ormond Street Hospital, Children's Hospital of Los Angeles, Children's Hospital of Philadelphia, Columbia University Medical Center, Icahn School of Medicine at Mount Sinai, Rochester School of Medicine and Dentistry, Steven and Alexandra Cohen Children's Medical Center of New York, and Yale School of Medicine approved the protocols. All subjects or their parents provided informed consent. Kids First project represents a subset of the PCGC data so CHD subjects were recruited as per PCGC study protocol.

