## Supplementary figures for "Genomic rare variant mechanisms for congenital cardiac laterality defect: A digenic model approach"

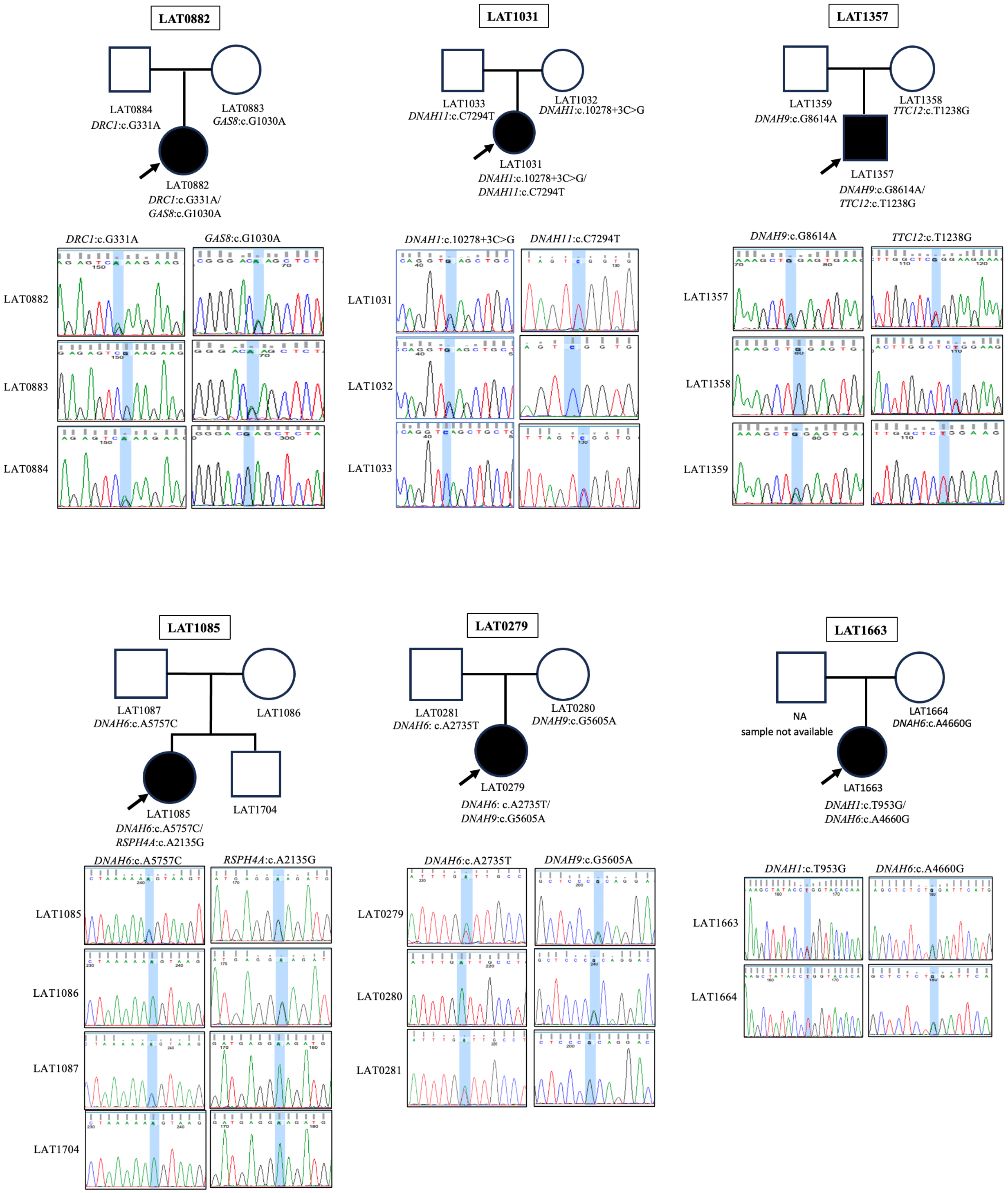
LAT0882
LAT0884
DRC1:c.G331A
LAT0883
GAS8:c.G1030A
LAT0882
DRC1:c.G331A/
GAS8:c.G1030A
LAT1031
LAT1033
DNAH11:c.C7294T
LAT1032
DNAH1:c.10278+3C>G
LAT1031
DNAH1:c.10278+3C>G/
DNAH11:c.C7294T
LAT1357
LAT1359
DNAH9:c.G8614A
LAT1358
TTC12:c.T1238G
LAT1357
DNAH9:c.G8614A/
TTC12:c.T1238G
DRC1:c.G331A
GAS8:c.G1030A
LAT0882
LAT0883
LAT0884
DNAH1:c.10278+3C>G
DNAH11:c.C7294T
LAT1031
LAT1032
LAT1033
DNAH9:c.G8614A
TTC12:c.T1238G
LAT1357
LAT1358
LAT1359
LAT1085
LAT1087
DNAH6:c.A5757C
LAT1086
LAT1085
DNAH6:c.A5757C/
RSPH4A:c.A2135G
LAT1704
LAT0279
LAT0281
DNAH6: c.A2735T
LAT0280
DNAH9:c.G5605A
LAT0279
DNAH6: c.A2735T/
DNAH9:c.G5605A
LAT1663
NA
sample not available
LAT1664
DNAH6:c.A4660G
LAT1663
DNAH1:c.T953G/
DNAH6:c.A4660G
DNAH6:c.A5757C
RSPH4A:c.A2135G
LAT1085
LAT1086
LAT1087
LAT1704
DNAH6:c.A2735T
DNAH9:c.G5605A
LAT0279
LAT0280
LAT0281
DNAH1:c.T953G
DNAH6:c.A4660G
LAT1663
LAT1664

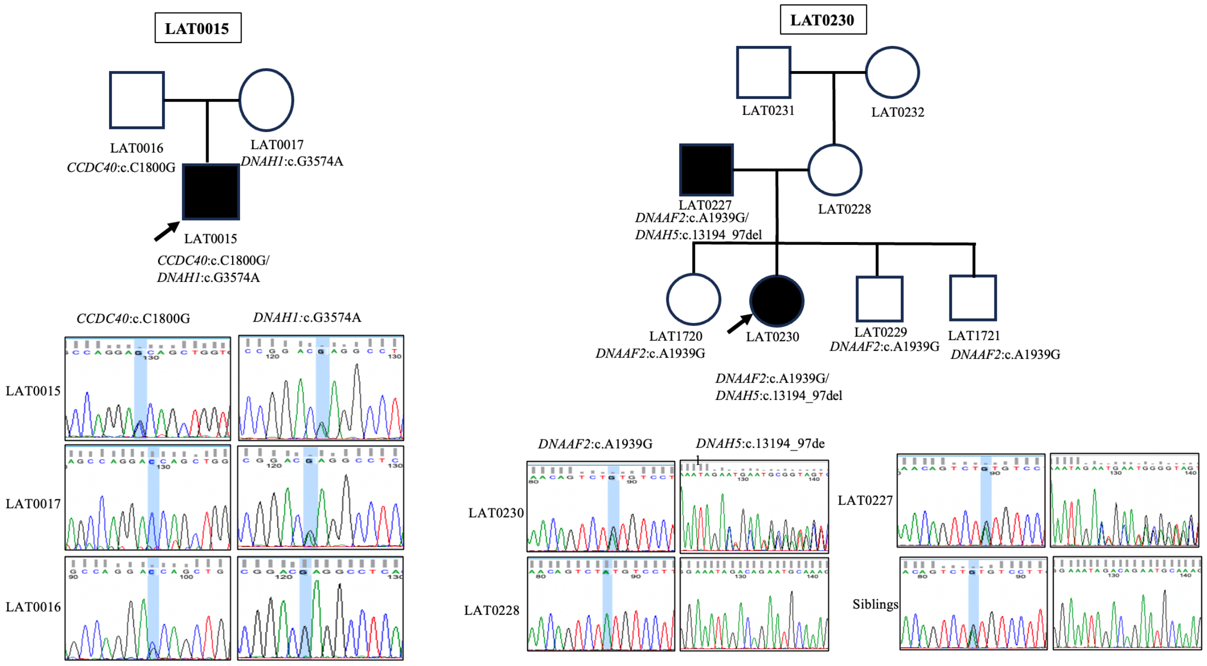


**Figure S1: Comprehensive pedigrees and their genotypes for families with digenic variants.** Standard pedigree structures are utilized – filled circles and squares denote clinically affected individuals. Lower panels indicate the Sanger sequencing data for available members of each family.

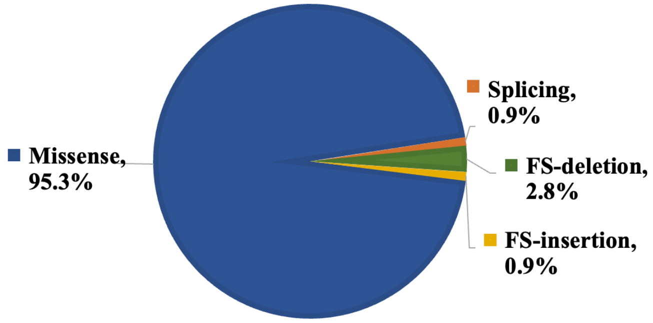


| Mutation type | Number of mutations |
|---|---|
| Missense | 101 |
| Splicing | 1 |
| FS-deletion | 3 |
| FS-insertion | 1 |

**Figure S2: The distribution of single nucleotide variant (SNV) types in Laterality Defect cases.** The pie chart shows the percentage of mutation types by color out of the total number of identified SNVs. Missense variants accounted for the majority (95.3%) of variants identified in three cohorts, followed by frameshift deletion (Fsdel, 2.8%), frameshift insertion (Fsins, 0.9%) and splice site variants (0.9%).
